# Cervical Cancer Screening with Visual Inspection with Acetic Acid (VIA) among Women Living with HIV in Nairobi, Kenya: Results and Transformation Zone

**DOI:** 10.64898/2026.03.27.26349550

**Authors:** Xinyi Feng, Ramya Ginjupalli, Judith Lukhorito, David Karanja, Mark Mounir, Mary Nderitu, Millicent Masinde, Sue Siminski, Lu Mao, Vikrant V. Sahasrabuddhe, Nazha M. Diwan, Michael H. Chung

**Author notes:** Corresponding Author*: Michael H. Chung, MD, Ph.D., MPH, Emory School of Medicine and Rollins School of Public Health, 80 Jesse Hill Jr Dr SE, Atlanta, GA 30303.

## Abstract

**Background:** Cervical cancer remains a major public health challenge among women living with HIV (WLWH) in sub-Saharan Africa, where screening coverage remains suboptimal despite opportunities for integration within HIV care programs. Visual inspection with acetic acid (VIA) has been widely used as a low-cost screening approach in resource-limited settings.

**Methods:** This cross-sectional analysis utilized prospectively collected data from Project CN001 at the Coptic Hope Center for Infectious Diseases in Nairobi, Kenya, a CASCADE Clinical Trials Network site. WLWH aged 25-49 years receiving routine HIV care and undergoing VIA screening between March 11, 2025, and January 16, 2026, were included. Data from the REDCap and Kenya’s electronic medical record system (KenyaEMR) captured sociodemographic characteristics, HIV clinical factors, VIA results, and cervical transformation zone (TZ) classification.

**Results:** Among 857 WLWH screened with VIA, the median age was 40 years (interquartile ranges [IQR]: 34-45), and 77.2% reported a prior history of cervical cancer screening. VIA positivity was 7.4% (63/857) and was higher in women with TZ1/TZ2 than in those with TZ3. VIA positivity was also associated with higher HIV viral load, shorter time since HIV diagnosis, no cervical screening history, and younger age at screening. The proportion of women classified as TZ3 increased with age, from 39.5% among women aged 25-29 years to 67.7% among those aged 45-49 years, while the proportion classified as TZ1 decreased with increasing age.

**Conclusion:** Integrated screening at this urban U.S. President’s Emergency Plan for AIDS Relief (PEPFAR) and CASCADE-supported HIV clinic demonstrates the feasibility of integrated cervical cancer screening programs for WLWH. Age-related TZ3 predominance and VIA limitations for older women highlight the need for refined screening strategies and continued electronic platform utilization for program monitoring to support cervical cancer elimination targets.

## Introduction

Cervical cancer remains a major global public health challenge and is disproportionately concentrated in low- and middle-income countries (LMICs), particularly in sub-Saharan Africa, where more than 85% of global cervical cancer deaths occur.^1^ Compared with women without HIV, women living with HIV (WLWH) have been estimated to have approximately six-fold higher risk of developing cervical cancer, making cervical cancer prevention a critical priority within HIV care programs.^2^

In many resource-limited settings, visual inspection with acetic acid (VIA) has been widely used for screening, allowing eligible women to receive immediate ablative treatment during the same visit.^3^ In Kenya, lifetime screening coverage among women aged 30-49 years was estimated at 21% in 2020, higher than the Eastern Africa regional average but still far below targets needed for effective cervical cancer prevention.^4^ Among WLWH, screening coverage has been reported to be even lower, with some studies suggesting fewer than 10% in high-burden settings.^5^

The U.S. President’s Emergency Plan for AIDS Relief (PEPFAR), through initiatives such as the Go Further partnership established in 2018, has supported the integration of cervical cancer screening using VIA into HIV treatment facilities across sub-Saharan Africa.^6,7^ The CASCADE Clinical Trials Network, funded by the U.S. National Cancer Institute (NCI), was established to improve cervical cancer screening, management, and precancer treatment among WLWH in high-burden settings.^8^

Project CN001, a component of the CASCADE network, was designed to establish a cervical cancer data repository to longitudinally track clinical outcomes of screening and precancer treatment in LMICs. The Coptic Hope Center for Infectious Diseases is one of the CN001 clinical sites where this database is implemented to: (1) evaluate the effectiveness of integrating cervical cancer screening and precancer treatment into HIV care systems by identifying gaps in screening uptake, diagnostic accuracy, and follow-up care; and (2) establish a robust infrastructure to facilitate recruitment of eligible participants into cervical cancer clinical trials assessing novel screening methods and treatment strategies.

Understanding the screening program, participant profiles, and screening outcomes is critical for monitoring implementation, assessing screening quality, and informing strategies to strengthen cervical cancer prevention among WLWH in high-burden settings. Therefore, in this study, we described the sociodemographic and clinical characteristics, and cervical cancer screening results among WLWH in Project CN001 who underwent screening with VIA, and evaluated factors associated with screening outcomes.

## Methods

### Study settings and population

The CASCADE Clinical Trials Network is an NCI-supported initiative aimed at improving cervical cancer screening, management, and treatment among WLWH in high HIV-burden settings. The network conducts pragmatic clinical and implementation research across multiple sites in sub-Saharan Africa and the United States to evaluate strategies for optimizing the cervical cancer prevention cascade among WLWH.^8^

This study was conducted at the Coptic Hope Center for Infectious Diseases in Nairobi, Kenya, a clinical site within the CASCADE network. Established in 2004, the Hope Center is a large HIV treatment facility providing care to over 54,000 adults and children, approximately 65% of whom are women. Cervical cancer prevention services at the Hope Center are integrated into routine HIV care and supported through PEPFAR via the Go Further partnership. Screening is primarily conducted during routine HIV clinic visits using VIA, performed by trained clinical providers (nurses) following standard procedures. During pelvic examination, a speculum is inserted and 5% acetic acid is applied to the cervix, which is then examined for acetowhite lesions suggestive of cervical precancer.

Project CN001 is a prospective clinical data repository of WLWH aged 25–49 years receiving care at clinical sites like the Coptic Hope Center. The repository was designed to longitudinally follow women through the cervical cancer prevention cascade, improve follow-up and retention in care, and facilitate identification and enrollment of eligible participants into CASCADE clinical trials. It captures data on screening, diagnostic evaluation, referral, and treatment, supporting ongoing monitoring of cervical cancer screening outcomes and program implementation.

This study included WLWH aged 25–49 years, consistent with Kenyan cervical cancer screening guidelines,^9^ who received HIV care at the Coptic Hope Center and underwent VIA screening between March 11, 2025, and January 16, 2026. Women with a history of cervical cancer or prior total hysterectomy were excluded.

Cervical cancer screening data, including age at screening, VIA results, transformation zone (TZ) classification, and prior screening history, were collected and managed using REDCap, with TZ classified as TZ1, TZ2, or TZ3. Date of HIV diagnosis and HIV-related clinical data closest to the time of cervical screening, including CD4 count, HIV viral load, and antiretroviral therapy (ART) regimen, were obtained from KenyaEMR. The lower limit of HIV viral load detection was defined as HIV RNA <50 copies/mL. Sociodemographic characteristics, including educational attainment, occupation, and marital status, were also obtained from KenyaEMR at the time of enrollment.

### Statistical Methods

Participant characteristics were summarized using descriptive statistics. Continuous variables were reported as medians with interquartile ranges (IQR), and categorical variables as frequencies and percentages. We examined the distribution of sociodemographic and clinical characteristics among women who underwent VIA screening, stratified by VIA results. Differences between VIA-positive and VIA-negative women were assessed using chi-square tests or Fisher’s exact tests for categorical variables, and Wilcoxon rank-sum tests for continuous variables due to skewed distributions. We additionally described the distribution of TZ classification across age groups. Statistical significance was defined as p < 0.05. All analyses were conducted using Stata version 15.1 (StataCorp, College Station, TX) and R version 4.2.1 (R Core Team, 2022).

### Ethics

This study used de-identified clinical data collected as part of the CASCADE cervical cancer screening program and Project CN001 database. The protocol was reviewed and approved by the Kenyatta National Hospital-University of Nairobi Ethics and Research Committee (Approval Ref Number P532/07/2024) and by the National Commission for Science, Technology, and Innovation in Kenya, as well as by the Emory University Institutional Review Board (IRB #STUDY00008254). All participants provided informed consent for collection and use of their clinical data. Data management procedures complied with institutional and regulatory requirements to ensure participant privacy and confidentiality.

## Results

### VIA screening results

Among 886 WLWH aged 25–49 years who were offered VIA screening, 857 (96.7%) underwent screening and were included in this study. Among those screened, 63 (7.4%) were VIA-positive, including 4 suspicious of cancer, while 798 (93.1%) were VIA-negative.

Table 1 summarizes the sociodemographic characteristics of the study population by VIA result. The median age of participants was 40 years (IQR: 34–45). Women with VIA-positive results were younger than those with VIA-negative results, with a median age of 35 years (IQR: 29–42) compared with 40 years (IQR: 34–45), respectively (*p* < 0.001). Educational attainment did not differ significantly by VIA result (*p* = 0.275). Overall, most women had completed secondary school (46.1%), followed by college, university, or polytechnic education (38.7%), and primary school education (14.5%). The most common occupation was trader (36.6%), followed by employee (28.6%), student or education (15.4%), and no occupation (11.5%). Marital status was also similar between groups (*p* = 0.726), with the majority of women married or in union (68.2%), followed by never married (18.6%) and previously married (8.0%).

**Table 1.**
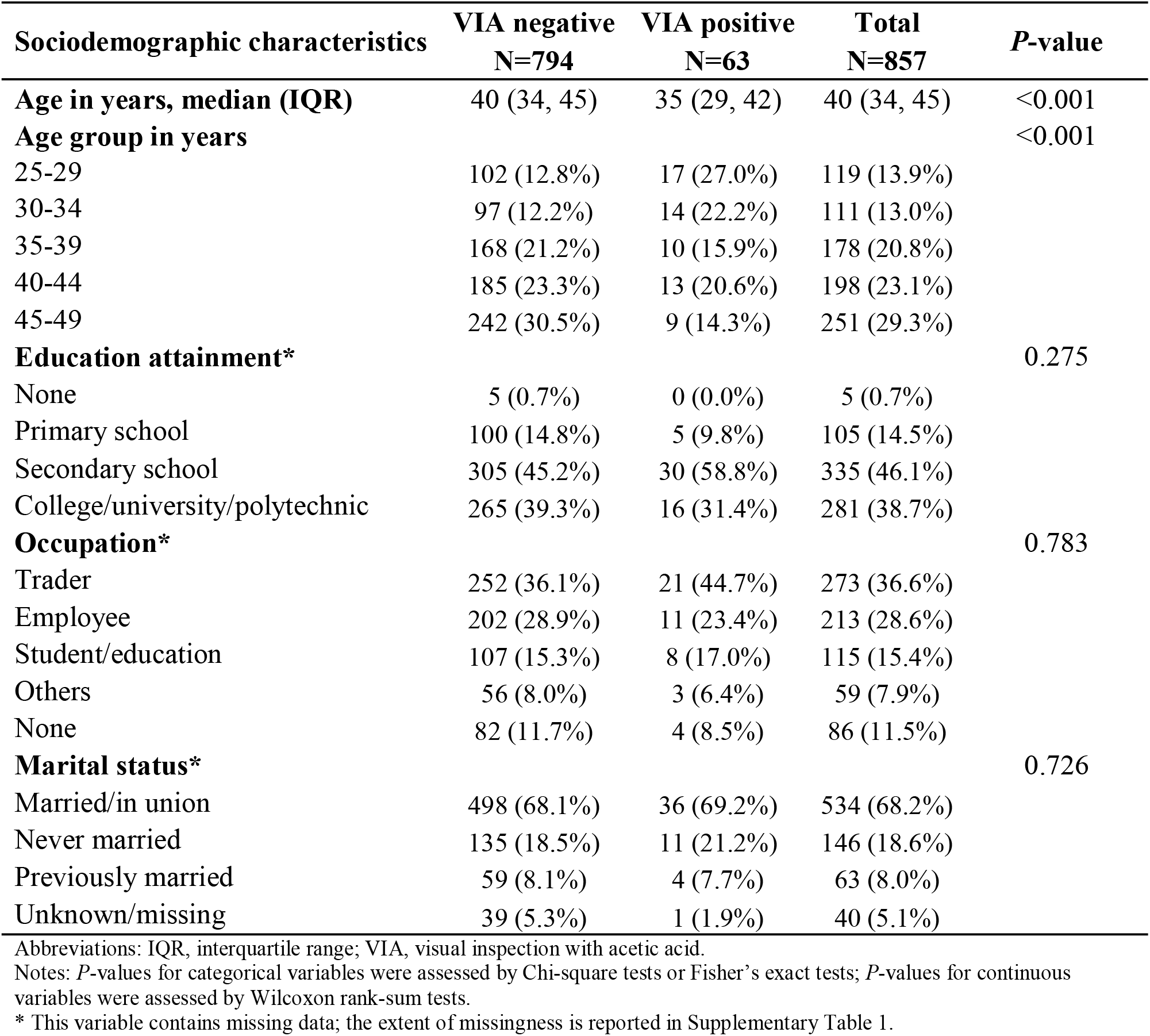
Sociodemographic characteristics of women living with HIV seen at the Coptic Hope Center for Infectious Disease in Nairobi, Kenya from March 11, 2025, to January 16, 2026 (N=857)

Table 2 presents the clinical characteristics stratified by VIA result. Women with VIA-positive results had a shorter duration since HIV diagnosis than VIA-negative women, with a median of 6 years (IQR: 3–13) compared with 10 years (IQR: 5–15) (*p* = 0.003). Most women were receiving the antiretroviral therapy regimen TDF/3TC/DTG (96.8%), and ART regimen did not differ significantly by VIA result (*p* = 0.270). CD4 count categories were similar between groups (*p* = 0.628), with 19.6% having <200 cells/µL and 42.5% having ≥500 cells/µL overall.

**Table 2.**
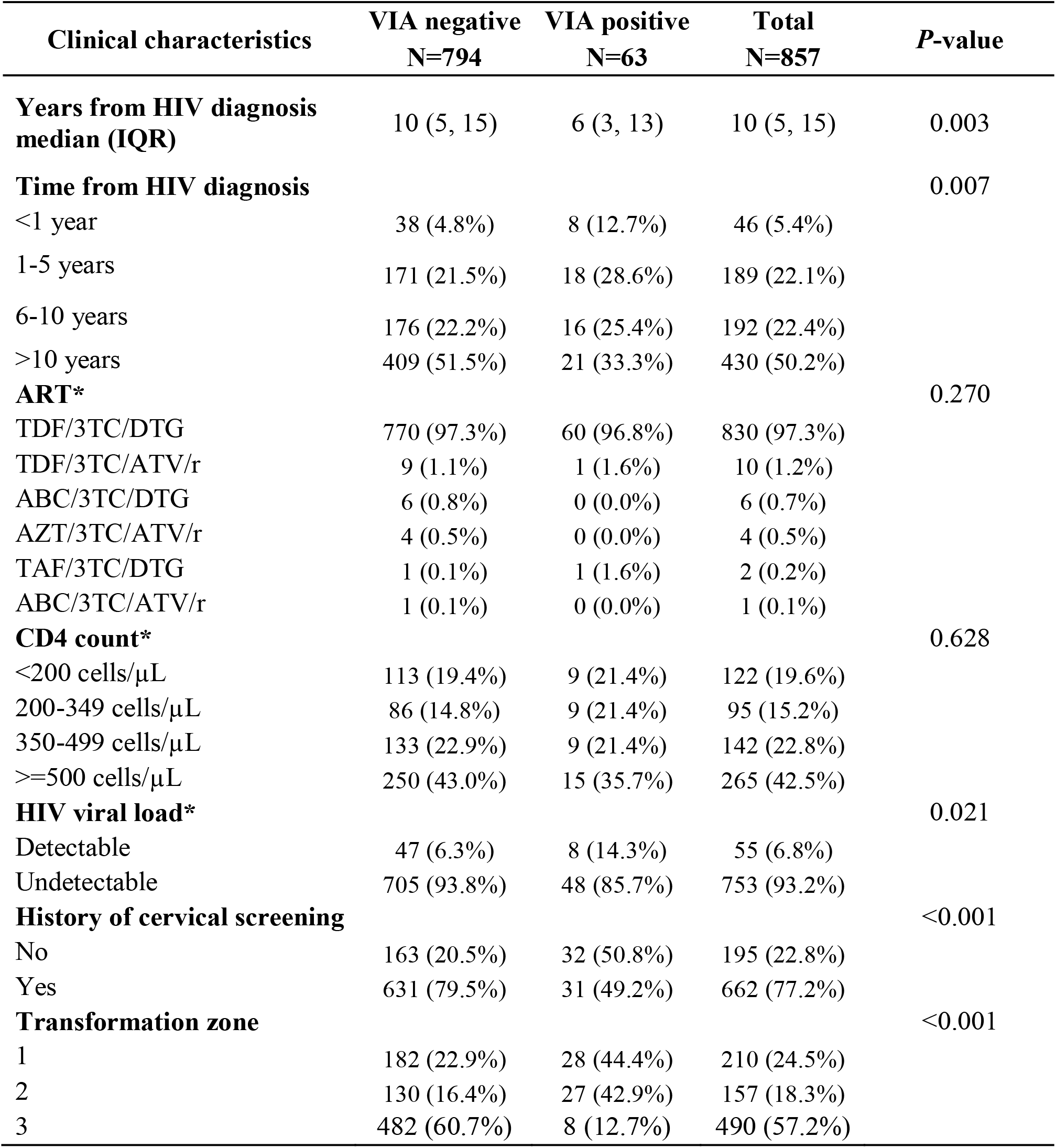
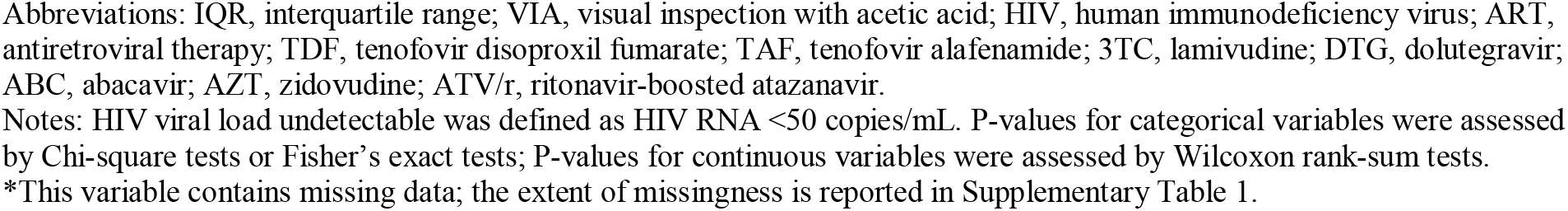
Clinical characteristics of women living with HIV seen at the Coptic Hope Center for Infectious Disease in Nairobi, Kenya from March 11, 2025, to January 16, 2026 (N=857)

| Clinical characteristics | VIA negative<br>N=794 | VIA positive<br>N=63 | Total<br>N=857 | P-value |
| --- | --- | --- | --- | --- |
| **Years from HIV diagnosis median (IQR)** | 10 (5, 15) | 6 (3, 13) | 10 (5, 15) | 0.003 |
| **Time from HIV diagnosis** |  |  |  | 0.007 |
| <1 year | 38 (4.8%) | 8 (12.7%) | 46 (5.4%) |  |
| 1-5 years | 171 (21.5%) | 18 (28.6%) | 189 (22.1%) |  |
| 6-10 years | 176 (22.2%) | 16 (25.4%) | 192 (22.4%) |  |
| >10 years | 409 (51.5%) | 21 (33.3%) | 430 (50.2%) |  |
| **ART\*** |  |  |  | 0.270 |
| TDF/3TC/DTG | 770 (97.3%) | 60 (96.8%) | 830 (97.3%) |  |
| TDF/3TC/ATV/r | 9 (1.1%) | 1 (1.6%) | 10 (1.2%) |  |
| ABC/3TC/DTG | 6 (0.8%) | 0 (0.0%) | 6 (0.7%) |  |
| AZT/3TC/ATV/r | 4 (0.5%) | 0 (0.0%) | 4 (0.5%) |  |
| TAF/3TC/DTG | 1 (0.1%) | 1 (1.6%) | 2 (0.2%) |  |
| ABC/3TC/ATV/r | 1 (0.1%) | 0 (0.0%) | 1 (0.1%) |  |
| **CD4 count\*** |  |  |  | 0.628 |
| <200 cells/μL | 113 (19.4%) | 9 (21.4%) | 122 (19.6%) | |
| 200-349 cells/μL | 86 (14.8%) | 9 (21.4%) | 95 (15.2%) | |
| 350-499 cells/μL | 133 (22.9%) | 9 (21.4%) | 142 (22.8%) | |
| ≥500 cells/μL | 250 (43.0%) | 15 (35.7%) | 265 (42.5%) | |
| **HIV viral load\*** |  |  |  | 0.021 |
| Detectable | 47 (6.3%) | 8 (14.3%) | 55 (6.8%) |  |
| Undetectable | 705 (93.8%) | 48 (85.7%) | 753 (93.2%) |  |
| **History of cervical screening** |  |  |  | <0.001 |
| No | 163 (20.5%) | 32 (50.8%) | 195 (22.8%) |  |
| Yes | 631 (79.5%) | 31 (49.2%) | 662 (77.2%) |  |
| **Transformation zone** |  |  |  | <0.001 |
| 1 | 182 (22.9%) | 28 (44.4%) | 210 (24.5%) |  |
| 2 | 130 (16.4%) | 27 (42.9%) | 157 (18.3%) |  |
| 3 | 482 (60.7%) | 8 (12.7%) | 490 (57.2%) |  |

Abbreviations: IQR, interquartile range; VIA, visual inspection with acetic acid; HIV, human immunodeficiency virus; ART, antiretroviral therapy; TDF, tenofovir disoproxil fumarate; TAF, tenofovir alafenamide; 3TC, lamivudine; DTG, dolutegravir; ABC, abacavir; AZT, zidovudine; ATV/r, ritonavir-boosted atazanavir.
Notes: HIV viral load undetectable was defined as HIV RNA <50 copies/mL. P-values for categorical variables were assessed by Chi-square tests or Fisher's exact tests; P-values for continuous variables were assessed by Wilcoxon rank-sum tests.
\*This variable contains missing data; the extent of missingness is reported in Supplementary Table 1.

In contrast, HIV viral load differed significantly by VIA positivity, with a higher proportion of detectable viral load among VIA-positive women (14.3%) compared with VIA-negative women (6.3%) (*p* = 0.021). A prior history of cervical cancer screening was less common among VIA-positive women (49.2%) than VIA-negative women (79.5%) (*p* < 0.001). Finally, TZ classification differed significantly by VIA result (*p* < 0.001), with VIA-positive women more likely to have TZ1 or TZ2.

### Distribution of transformation zone by age

The majority of WLWH in this study had TZ2 or TZ3 (>75%). Overall, TZ3 was the most common classification (490/857, 57.2%), followed by TZ1 (210/857, 24.5%) and TZ2 (157/857, 18.3%).

The distribution of TZ types varied substantially by age group (Table 3). Younger women were more likely to have TZ1, while older women were more likely to have TZ3. Among women aged 25–29 years, 41.2% had TZ1 compared with 19.3% who had TZ2, and 39.5 who had TZ3. In contrast, among women aged 45–49 years, 67.7% had TZ3 compared with 15.1% who had TZ1. The proportion of TZ3 increased steadily with age, while the proportion of TZ1 declined with increasing age.

**Table 3.** Distribution of cervical transformation zone classification by age group among women living with HIV seen at the Coptic Hope Center for Infectious Disease in Nairobi, Kenya from March 11, 2025, to January 16, 2026 (N=857)

| Age group | TZ1 | TZ2 | TZ3 | Total |
| --- | --- | --- | --- | --- |
| 25–29 | 49 (41.2%) | 23 (19.3%) | 47 (39.5%) | 119 |
| 30–34 | 44 (39.6%) | 24 (21.6%) | 43 (38.7%) | 111 |
| 35–39 | 52 (29.2%) | 30 (16.9%) | 96 (53.9%) | 178 |
| 40–44 | 27 (13.6%) | 37 (18.7%) | 134 (67.7%) | 198 |
| 45–49 | 38 (15.1%) | 43 (17.1%) | 170 (67.7%) | 251 |
| **Total** | **210** | **157** | **490** | **857** |

## Discussion

This study describes cervical cancer screening outcomes using VIA among 857 WLWH receiving routine HIV care at the Coptic Hope Center in Kenya, an urban PEPFAR-supported clinic within the CASCADE Clinical Trials Network. Overall, 7.4% of women screened were VIA-positive, which was associated with younger age, shorter time since HIV diagnosis, higher HIV viral load, lack of prior screening, and TZ classification. We also observed age-related differences in TZ types, with TZ3 increasing and TZ1 decreasing with age.

Among 857 women, 7.4% tested VIA-positive, a proportion slightly higher than the <5% positivity reported in many Kenyan programs but lower than the 15-20% positivity observed in large western Kenya HIV cohorts.^10,11^ These differences likely reflect both programmatic and population factors, including higher prior screening coverage and better viral suppression at this site, which may reduce the underlying burden of high-grade lesions while still enabling detection of prevalent abnormalities among women engaged in long-term ART.^12^

VIA positivity was associated with younger age, shorter time since HIV diagnosis, and higher viral load, consistent with prior HIV-focused screening studies.^14^ These findings are biologically plausible, as poor viral control and recent HIV infection are linked to increased HPV persistence and progression to cervical precancer. ^15^ In addition, these associations may reflect underlying socio-behavioral factors, including differences in sexual exposure and access to care, which influence both HIV control and cervical cancer risk.^16^

Notably, among 857 screened WLWH, 77.2% had documented prior screening, substantially exceeding coverage reported in other Kenyan HIV programs, where lifetime screening has been estimated at 29.4% and annual rates are typically below 10%.^10^ Enhanced urban infrastructure, robust PEPFAR support, and CASCADE’s documentation likely account for this disparity, addressing implementation challenges noted in prior Kenyan studies. Fewer documented prior screenings among VIA-positive women suggest under-detection in earlier VIA assessments or incomplete documentation, underscoring the importance of electronic health record integration for routine monitoring. However, these findings reflect an urban cohort with long-term ART engagement and high viral suppression, and may not generalize to younger WLWH, underserved populations (e.g., sex workers), or newly diagnosed individuals at higher risk of cervical disease.^17^

The high prevalence of TZ3, particularly among older women (67.7% among those aged 45–49 years), is consistent with age-related cervical remodeling following childbearing and declining estrogen levels, which limit visualization of the transformation zone.^18^ Among WLWH, prolonged survival on ART, cumulative HPV exposure, and prior cervical treatments may further accelerate this shift toward TZ3 and contribute to scarring or cervical stenosis that make complete visualization more difficult. Kenyan and regional data affirm VIA’s moderate sensitivity and variable specificity, contingent on provider expertise and TZ visibility.^13^ While limited studies have been shown to link predominance of TZ3 in older WLWH, this highlights the potential intersection of aging, HIV, and reproductive history in shaping transformation zone anatomy.^19^ Our findings thus align with broader regional data and underscore that TZ3 is not only a technical challenge but a predictable consequence of aging WLWH populations on ART.^18,20^ The predominance of TZ3 among older WLWH also underscores an important challenge: when the transformation zone is not fully visible, VIA performance may be compromised and additional or alternative triage strategies are needed for accurate detection of precancer among WLWH in their 40s, supporting WHO guidance for HPV-based primary screening in WLWH and women with TZ3.^21^

The integrated structure of the Coptic Hope Center, supported by the CASCADE network and the CN001 data repository, likely contributed to both the high coverage and quality of VIA screening observed in this study. PEPFAR funding enabled dedicated screening staff, reliable supplies, structured screening workflows, consistent provider training, and routine quality assurance, all of which may enhance screening performance compared with less-resourced settings.^13^. These programmatic advantages contrast with vertical, non-integrated programs in Kenya, where cervical cancer screening remains sporadic, coverage is low, and VIA positivity is often derived from short-term campaigns or targeted interventions rather than sustained service delivery.^10,22^ By embedding screening within routine HIV care and leveraging KenyaEMR and CN001 for ongoing monitoring of coverage, test results, and follow-up, this model enables more systematic identification of VIA-positive lesions that might otherwise be missed in less-resourced, stand-alone screening settings.

This study has several limitations. First, it was conducted within a single urban HIV treatment program, which may limit generalizability to rural settings and high-risk populations. Second, the analysis relied on routinely collected KenyaEMR data, and missingness in key sociodemographic and clinical variables may have introduced bias if not random. Third, the cross-sectional design precludes assessment of longitudinal outcomes, including lesion progression, treatment completion, and cervical cancer incidence. Finally, although we examined factors such as viral load and TZ type, unmeasured factors—including sexual behavior, prior screening outside the program, and provider-level variability in VIA interpretation—may have influenced screening outcomes.

However, by characterizing screening metrics within an integrated urban HIV–cervical cancer program, this study contributes a large sample of WLWH, addresses gaps in understanding cascade attrition in Kenya, and highlights VIA patterns that may inform more refined triage strategies. These findings demonstrate how CASCADE surveillance, integrated within KenyaEMR, can support routine monitoring of screening coverage, test positivity, and key risk indicators across PEPFAR-supported sites, and help track progress toward WHO cervical cancer elimination targets among WLWH.^23^

## Conclusion

Although cervical cancer screening coverage remains suboptimal across sub-Saharan Africa, our findings demonstrate the feasibility of integrating VIA screening within urban HIV care programs. High prior screening and strong engagement among women on long-term ART highlight the potential of PEPFAR-supported programs like CASCADE integration with electronic platforms such as KenyaEMR to reach WLWH, monitor screening quality, and identify high-risk subgroups. However, the predominance of TZ3 with increasing age underscores important limitations of VIA for older women. Continued integration of these data repository, alongside longitudinal CASCADE surveillance, will be essential for achieving WHO 90–70–90 cervical cancer elimination targets in high-burden settings.^23^

## Data Availability

All data produced in the present study are available upon reasonable request to the authors.

