## Supplementary Table 1 for "Cervical Cancer Screening with Visual Inspection with Acetic Acid (VIA) among Women Living with HIV in Nairobi, Kenya: Results and Transformation Zone"

**Supplementary Table 1.** Missingness of demographic and clinical variables by VIA result among women living with HIV seen at the Coptic Hope Center for Infectious Disease in Nairobi, Kenya from March 11, 2025, to January 16, 2026 (N=857)

| **Variable** | **VIA negative** | **VIA positive** | **Total** |
| --- | --- | --- | --- |
|  | **N=794** | **N=63** | **N=857** |
| **Education attainment** | 119 (15.0%) | 12 (19.0%) | 131 (15.3%) |
| **Occupation** | 95 (12.0%) | 16 (25.4%) | 111 (13.0%) |
| **Marital status** | 63 (7.9%) | 11 (17.5%) | 74 (8.6%) |
| **ART** | 3 (0.4%) | 1 (1.6%) | 4 (0.5%) |
| **CD4 count** | 212 (26.7%) | 21 (33.3%) | 233 (27.2%) |
| **HIV viral load** | 42 (5.3%) | 7 (11.1%) | 49 (5.7%) |
